# Genetic determination of regional connectivity in modelling the spread of COVID-19 outbreak for improved mitigation strategies

**DOI:** 10.1101/2021.01.30.21250785

**Authors:** Leonidas Salichos, Jonathan Warrell, Hannah Cevasco, Alvin Chung, Mark Gerstein

## Abstract

Covid-19 has resulted in the death of more than 1,500,000 individuals. Due to the pandemic’s severity, thousands of genomes have been sequenced and publicly stored with extensive records, an unprecedented amount of data for an outbreak in a single year. Simultaneously, prediction models offered region-specific and often contradicting results, while states or countries implemented mitigation strategies with little information on success, precision, or agreement with neighboring regions. Even though viral transmissions have been already documented in a historical and geographical context, few studies aimed to model geographic and temporal flow from viral sequence information. Here, using a case study of 7 states, we model the flow of the Covid-19 outbreak with respect to phylogenetic information, viral migration, inter- and intra-regional connectivity, epidemiologic and demographic characteristics. By assessing regional connectivity from genomic variants, we can significantly improve predictions in modeling the viral spread and intensity.

Contrary to previous results, our study shows that the vast majority of the first outbreak can be traced to very few lineages, despite the existence of multiple worldwide transmissions. Moreover, our results show that while the distance from hotspots is initially important, connectivity becomes increasingly significant as the virus establishes itself. Similarly, isolated local strategies-such as relying on herd immunity-can negatively impact neighboring states. Our work suggests that we can achieve more efficient unified mitigation strategies with selective interventions.

## INTRODUCTION

Covid-19 related deaths have surpassed 1,500,000 worldwide and 330,000 in the United States. Due to the importance of the pandemic, many resources are available for COVID-19 genome research, including GenBank, GISAID, and Nextstrain ^1–3^. Due to the severity of the pandemic combined with the advent of sequencing technologies, the amount of sequencing data within such a short time period for a single outbreak is unprecedented. GISAID is currently the largest COVID-19 database with more than 309,000 SARS-CoV2 genomes ^2^. This is compared to 1760 sequences of influenza A/H3N2 collected from 2013 to 2020. These numbers are comparable or surpassing the number of HIV or HCV sequences in the Los Alamos national database ^4,5^. These COVID-19 genomes represent the spread of the pandemic from China to 188 countries worldwide, with more sequences added every day.

Recent studies have modelled the transmission, diversity and spatial phylogeography of the virus mostly in a historical context ^6–12^. According to studies, Covid-19 first arrived in Washington ^11^, in what is considered a cryptic infection ^11,13^. However, known cases in persons with no relevant travel history also occurred in California in late January/early February^11^.While the first lineages arrived from China in Washington and California, subsequent infectious lineages (notably in New York) appear to represent importations from Europe ^11,14^. In this context, early results also suggested multiple worldwide transmissions responsible for the outbreak in the North-East of United States ^12^.

From the beginning of the pandemic, different approaches have been developed for the modeling of the outbreak that use either epidemiological, demographic data ^15–20^. Many-sometimes contradicting-prediction models offered temporal, and locally isolated results based on local outbreaks ^16–18,21,22^, while each country implemented their strategy to combat the outbreak, including controversial approaches such as “herd immunity”^23–25^. At the same time, different forms of local lockdowns have been tested to successfully mitigate viral spread as non-pharmaceutical interventions (NPIs) ^22,26–28^. While previous studies offer extremely valuable insights into the history of viral transmission and the effectiveness of locally implemented NPIs, they tend to overlook the inland spread (migration) of the virus in order to provide a unified mitigation strategy that complements local implementations.

In this study, we show a strong association between the temporal and geographical spread of the virus. By using a case study of seven states [New York (NY), New Jersey (NJ), Connecticut (CT), Massachusetts (MA), Pennsylvania (PA), Maryland (MD) and Virginia(VA)], we utilize the concepts of ingrowing, incoming and outgoing viral connectivity between states and regions as factors that influence the spread and viral transmission. We then use regression and random walk models to show the importance of these concepts combined with epidemiological and demographic factors-such as transmission rates and Urbanization Index in providing more informative predictions and explaining the temporal and geographic spread of the pandemic. The significance of modeling the spread of the viral wave through geographical routes and regional connectivity reveals broader implications and opportunities for the consideration of more efficient mitigation strategies in blocking viral migration with additional selective interventions.

### Initial Distance From Hotspots Determines Outbreak Severity

Using the numbers of ‘deaths per 1 million population’ as a proxy for regional outbreak severity, first we aimed to assess the association between distance from initial viral hotspots and the severity of the viral outbreak. To assign a geographic location for each state, we used the longitude and latitude from its respective larger city. Then, we considered the distance from New York City (New York), Seattle (Washington) and New Orleans (Louisiana) as the three initial hotspots of the outbreak. Introducing New York City (NYC) as a single initial hotspot, showed a high negative correlation (*r*=-0.37) between the severity of the outbreak and the distance from hotspot. By including Seattle (or San Francisco) as a second hotspot, the association increased (*r*=-0.43). Finally, by fitting a logarithmic curve, the association increased further to R^2^ =0.35 (figure 1). The inclusion of New Orleans as a third hotspot did not improve our results, indicating an isolated outbreak. On the contrary, by removing Louisiana as an outlier, we improved the predictability of the logarithmic curve to R^2^ =0.4. These results suggest a very high association between the outbreak’s severity during the first wave and the distance from the two initial hotspots.

**Figure 1.**
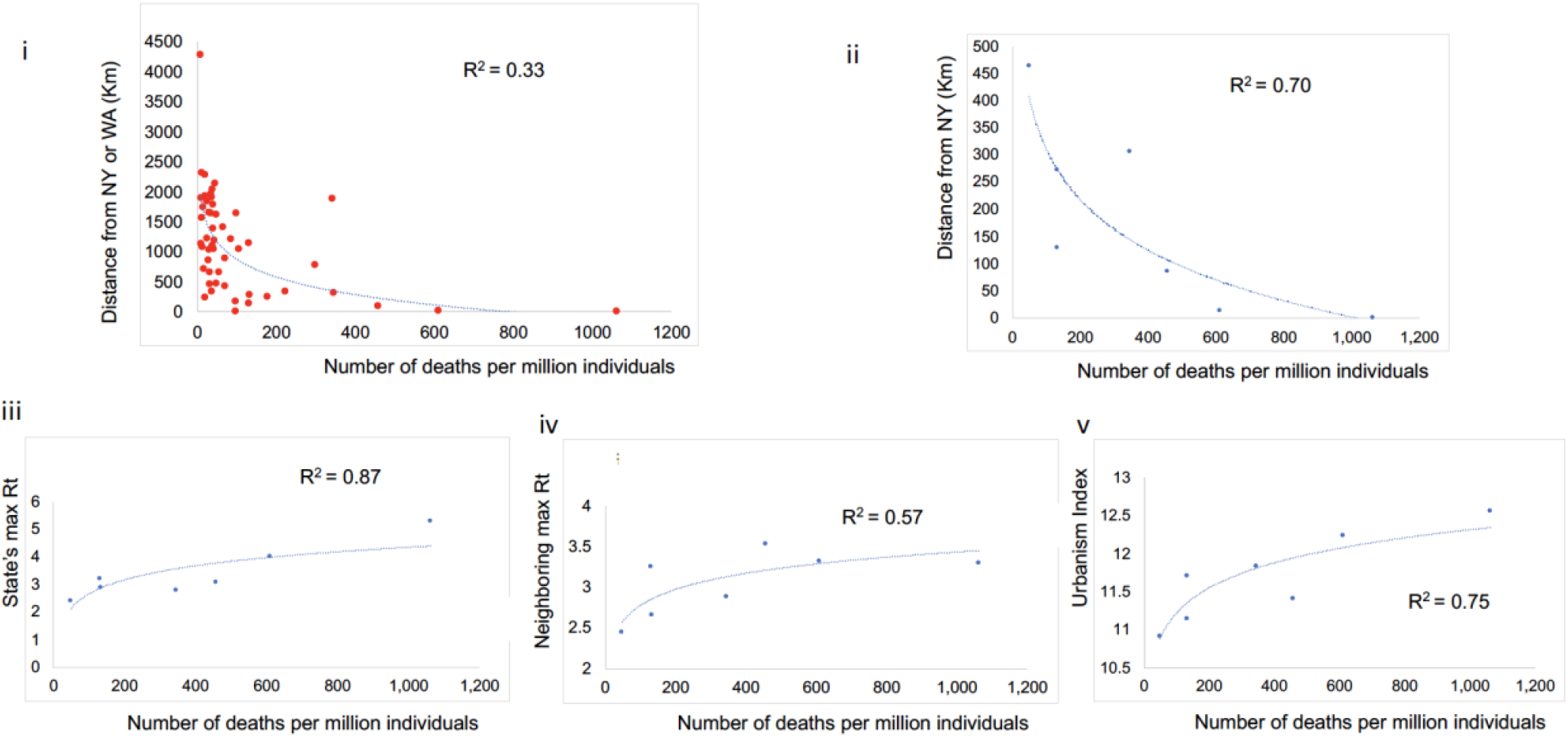
Using data collected on the 29th of April we show the logarithmic association between i) the number of deaths per million individuals for every state and the distance from hotspots (New York or Washington). Using a case study of 7 states (New York, New Jersey, Connecticut, Massachusetts, Pennsylvania, Virginia and Maryland), we show the logarithmic association between the number of deaths per million individuals versus ii) the Distance from New York city, iii) each state’s maximum reproduction rate Rt, iv) each state’s average neighboring maximum Rt, and v) each state’s urbanization index.

By fitting a log curve for the case study of seven states (NY, CT, MA, NJ, PA, MD and VA), we were able to associate the distance from NYC and the severity of the initial spread by explaining 70% of the variance. Additional factors strongly linked to the spread and severity of the epidemic include Urbanization Index and maximum effective reproduction rate R_t_ per state during the first wave, as retrieved from “https://rt.live/us/” (figure 1).

### Major 7-state Outbreak is Related to Few European Lineages

To create a dataset of world reference sequences, on June 25th, we sampled 50 sequences spanning the 5 Covid-19 lineages as determined by Nextstrain (Figure 2i). Lineages 19A and 19B represent the earliest detected infections closely associated with the Wuhan epidemic. Using the GISAID database, we downloaded all available sequences for our 7 states case study until the 6^th^ of August 2020. These sequences represent the first viral wave in the USA. Then, we inferred a phylogenetic tree for each individual state using Bayesian inference with date constraints under a Yule process population model. For all states, the major outbreak clusters with a specific European lineage (reference sequence: HF1465 FRA). For New York, Connecticut and Massachusetts this lineage clearly constitutes the dominant outbreak. For New Jersey, Pennsylvania, Maryland and Virginia, a secondary outbreak-which also circulates in NY, CT and MA- appears significant (figure 2, S1-7).

**Figure 2.**
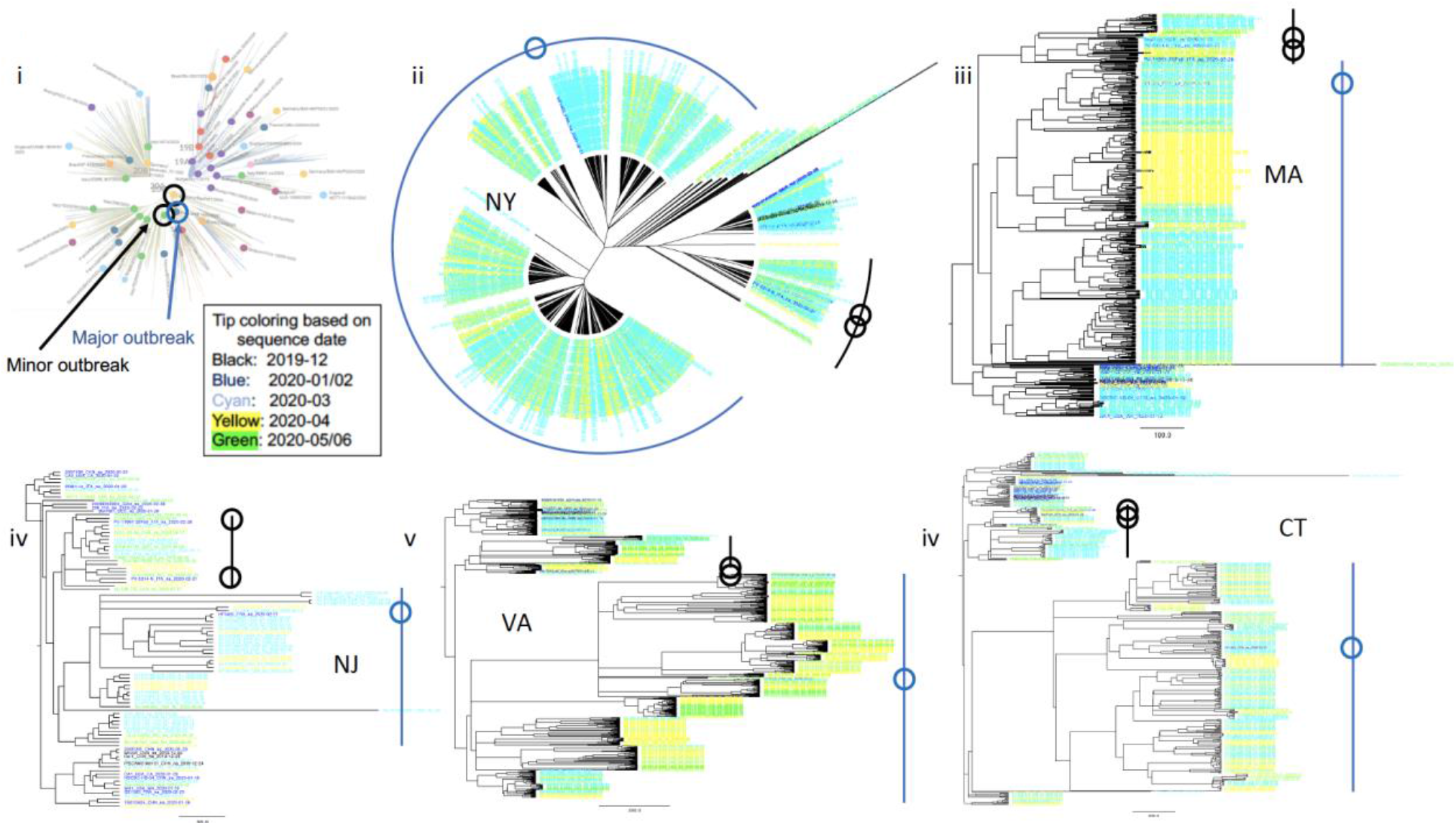
According to a Nextstrain adaptation, there were five main initial lineages of the pandemic (19A, 19B, 20A, 20B, 20C), which can be used to suggest the original routes of the transmission in the United States. In 2i) we show the topology of world reference sequences as collected spanning the Nextstrain tree on June 25th. From these randomly collected sequences, sample HF1465 FRA is the only sequence that consistently clusters with each state’s major outbreak (blue line). Two other reference sequences (from Italy and Germany) cluster-again consistently-with each state’s minor outbreak (black dotted line), suggesting that most of the outbreak derives from these specific lineages. In (2ii) we show the unrooted tree of the New York outbreak, which we consider as the outbreak epicenter. In (2iii-vi) we show the phylogenetic tree analysis for Massachusetts, New Jersey, Virginia and Connecticut as rooted by the older lineage that contains sequences from Wuhan dating in 2019.

### Assessing Viral Connectivity Between States

By selecting a set of (whenever possible) 50 reference sequences per state (in addition to the set of world reference sequences), we built a phylogenetic tree that includes sequences from all 7 states. We then inferred a connectivity map between the different states by parsing the tree’s bipartitions (figure 3i). For this, we examined all possible connected pairs of sequences that cluster together, while moving hierarchically from smaller to larger bipartitions without double-counting. To establish directionality between pairs, we used sampling dates. For example, the pair (NY-PV09151_USA_NY_2020-03-22, CT-UW-6574_USA_CT_2020-04-03) would be counted as NY -> CT, which denotes one incoming transitional connectivity from NY to CT. Similarly, the pair (NY-PV08434_USA_NY_2020-03-18, NY-NYUMC659_USA_NY_2020- 03-18) would be counted as ingrown connectivity NY -> NY. Overall, the NY outbreak showed the highest connectivity, while VA and MA showed the lowest. Interestingly, even though CT showed high connectivity comparable to NJ, the decreased number of outgoing versus incoming connections explains the low connectivity shown by MA. This is also supported by the outbreak’s high transitional connectivity from NY to CT (NY ->CT) rendering CT as a potential bottleneck (figure 3).

**Figure 3.**
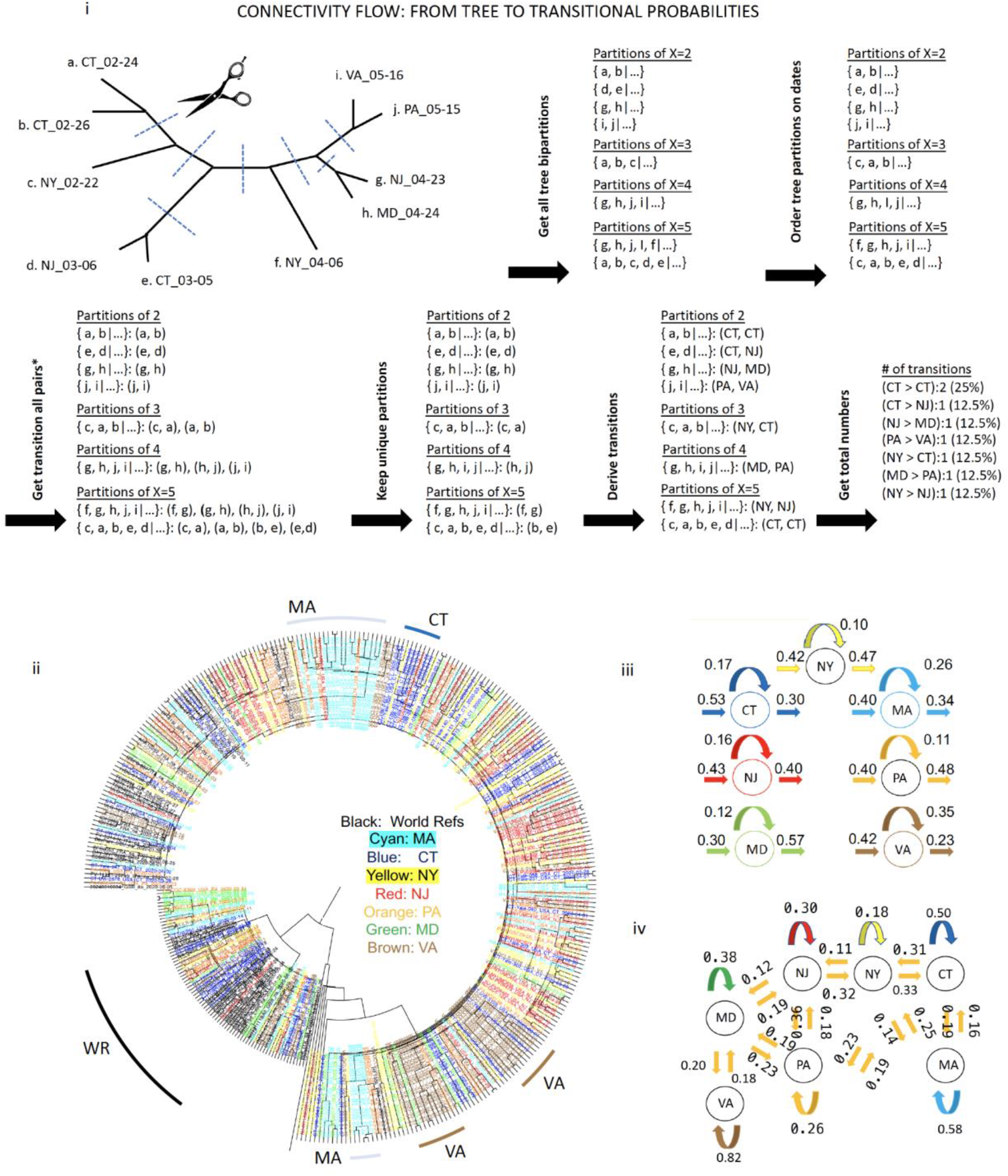
In i) we use a tree adaptation example to explain the workflow that we implemented in order to assign directed connectivity, incoming, ingrowing and outgoing connections between each state. In ii) using world reference sequences and selected reference sequences from 7 states, we inferred a phylogenetic tree with time constraints for each state. Each sequence’s tip color corresponds to the state it was collected. Using pairing and dating information described in (i), we derived iii) incoming, outgoing and ingrowing connectivity for each state and iii) transitional connectivity between all states. For convenience, we only show neighboring and geographical connectivity.

### Urbanism and Transitional Connectivity Increase Outbreak’s Severity

Previously, by considering the geographic distance from the initial hotspots NY and WA, we found a strong association between the distance and the severity of the outbreak for the first month of the epidemic. Additional factors associated with the spread included urbanism, the maximum R_t_ per state, as well as the neighboring states’ maximum R_t_. Here, we test the importance of various features in predicting the per-state death rate across the first wave of the pandemic (March to August 2020). We include in our analysis features including the estimated incoming, outgoing and ingrowing transmission rates between states, and a transmission-based normalized distance of each state from New York representing the viral flow (described below). The full feature set includes: maximum Reproduction rate per state (Rt) usually in April, Urbanization Index (U), Geographic or transition-based distance from New York (D or trD), and incoming, outgoing and ingrowing transmission rates. To determine the importance of regional transitional connectivity in addition to these factors in explaining and predicting the outbreak intensity during the whole first wave, we built 4 regression models with increasing complexity that combine phylogenetic information with epidemiological data from 10 dates (April 29^th^, May 1^st^ 8^th^ and 15^th^, June 11^th^ 18^th^ 25^th^, July 2^nd^, 29^th^, August 23^rd^).

In our simplest model (figure 4i) we examined the role of urbanism, distance (D) from NYC and virus’ maximum reproduction rate R_t_. U and D showed high and increasing significance throughout the whole first wave, while the use of maximum R_t_-as obtained by ‘https://rt.live/us/’- showed maximum significance at the beginning of the outbreak but eventually decreased. This is possibly because max R_t_ only represents the virus’ reproductive rate during the first stage of the outbreak (i.e in March-April), before the lockdowns. In our second model (figure 4ii), we substituted D with Transitional Distance (trD), a weighted version of distance as a proxy for viral flow which considers transitional connectivity between states (e.g. NY -> CT, NY -> NY, CT -> NY, NY -> NJ, .. etc) using random walks between the states (see methods). By replacing D with trD we were able to significantly increase our model’s predictability throughout the first wave (p=0.0003, figure S8). In our third model (figure 4iii), we returned to using the geographic distance D, but this time we also included each states’ total incoming, outgoing and ingrowing rates. Finally, in our 4^th^ model (figure 4iv), we again replaced D with trD, while also including states’ incoming and outgoing rates. While our 4^th^ model also integrates transitional connectivity in trD, this information is also used in calculating each state’s incoming, outgoing and ingrowing connectivity. Therefore, as expected, factors trD, incoming and outgoing rates often behave in a complementary manner. However, model 4 is still significantly more informative than model 3 (p=0.0273). Moreover, model 4 indicates that the initial importance of trD during the beginning of the outbreak, is gradually replaced by the state’s connectivity rate, as the outbreak spreads away from the initial hotspots.

**Figure 4.**
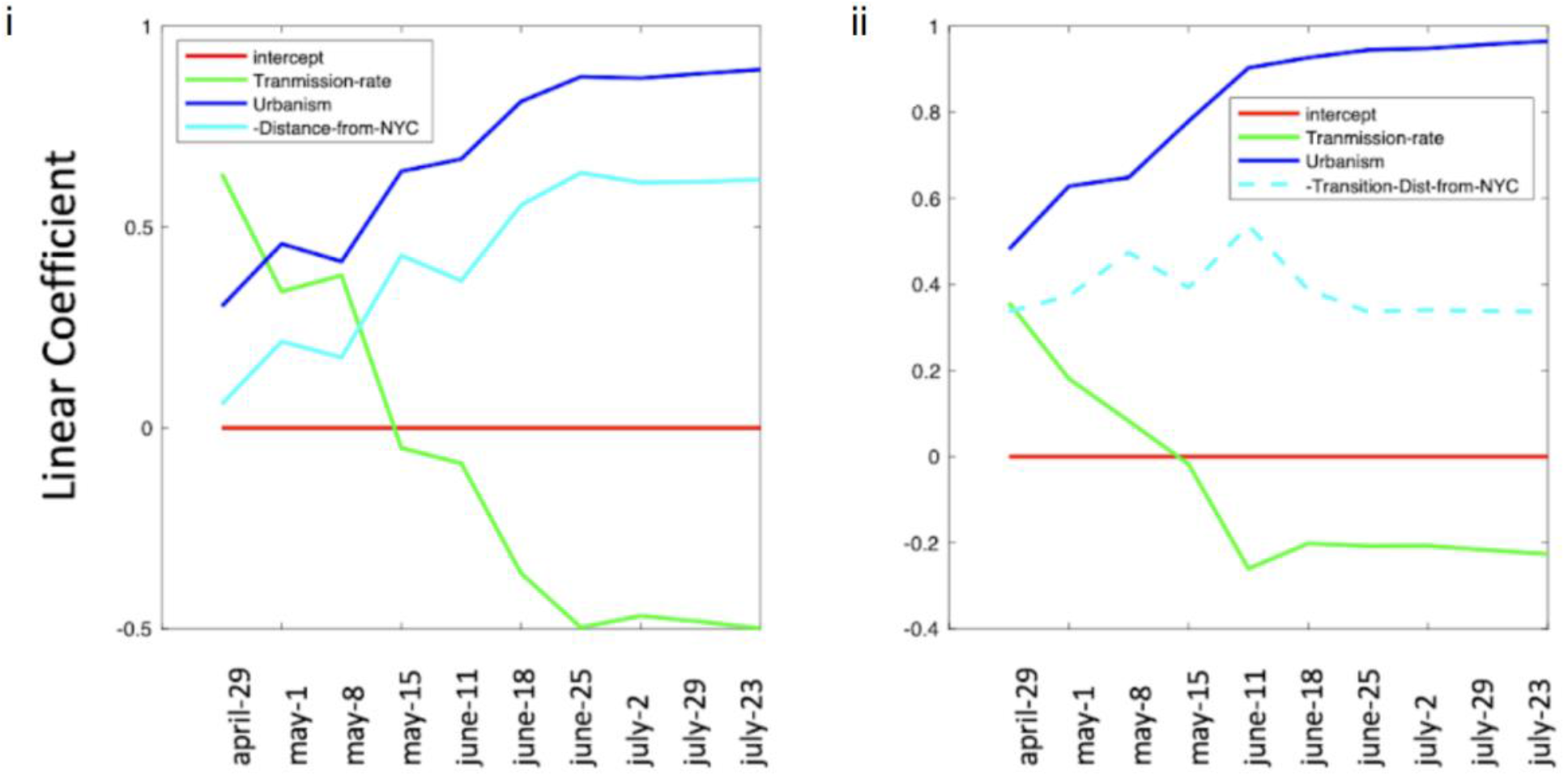
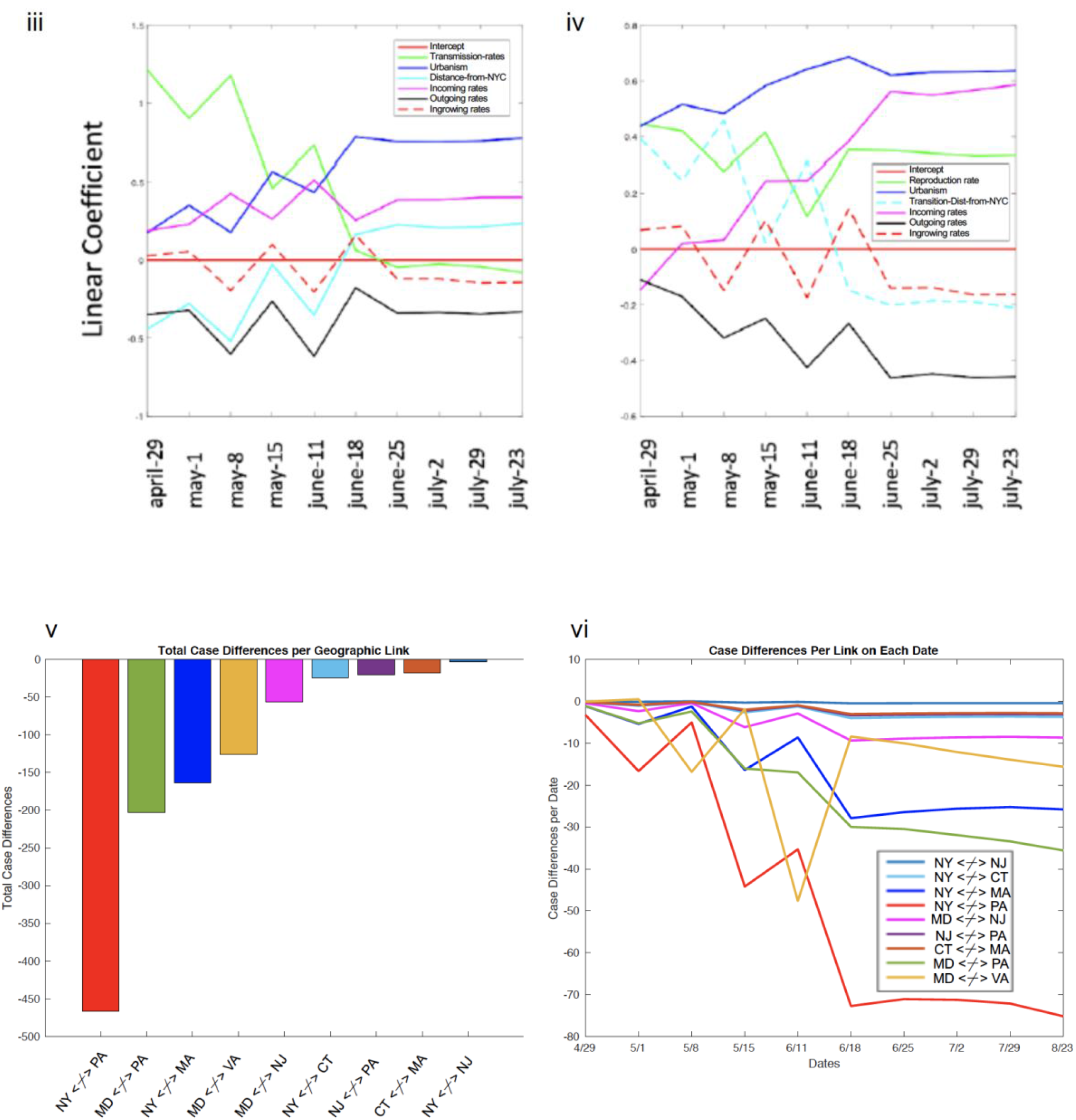
Predictive models with connectivity-based features. (i-ii) Models 1-2 (three factors), (iii-iv) Models 3-4 (six factors). Likelihood significance was found for models (1) vs (2) and (3) versus (4). (Model 1 vs. 2 / 3 vs. 4; p=0.0003, 0.0273 resp., 2-sided t-test for Pearson’s r). We then estimated the sum of total deaths that would be saved if we remove any geographic link between two states. In v) we show the total number of deaths per million individuals per case, while in vi) we show the temporal distribution of these deaths, showing when specific links become important. The link between NY and PA becomes important around May, while the link between MD and PA a month later.

### A Case Study for Selective Mitigation Strategies Based on Regional Connectivity

Using our second regression model with normalized transitional distance trD, we predicted the total number of deaths by removing one by one each geographic connection between every geographically linked state pair according to figure 3iv. Our results suggested that by enforcing a blockade between New York and Pennsylvania, as well as between Maryland and Pennsylvania would result in saving around 450 and 200 deaths per million individuals respectively, after the lockdowns. This is a particularly interesting result, since our model seems to take into account the drop in deaths in specific states after the imposed lockdowns (based on epidemiological data from NJ, NY, MA and CT) and respond to the temporal flow of the pandemic resulting in later death peaks in states like Virginia (figure S9). This becomes more evident in figure 4vi, where we depict the temporal effect of each blockade in reducing the number of total deaths per million individuals.

### New York’s Second Wave

To understand the origin of New York’s second wave, we inferred a new phylogenetic tree, this time by including all sequences from GISAID after August 2020 and until November 24th (figure 5). In addition to these new sequences, we also included our set of 50 world reference sequences and 50 from NY as mentioned previously. Our results indicate that about half of the second NY outbreak, has been re-introduced from Europe, possibly from Great Britain. This appears to be a completely new lineage, previously unseen in New York.

**Figure 5.**
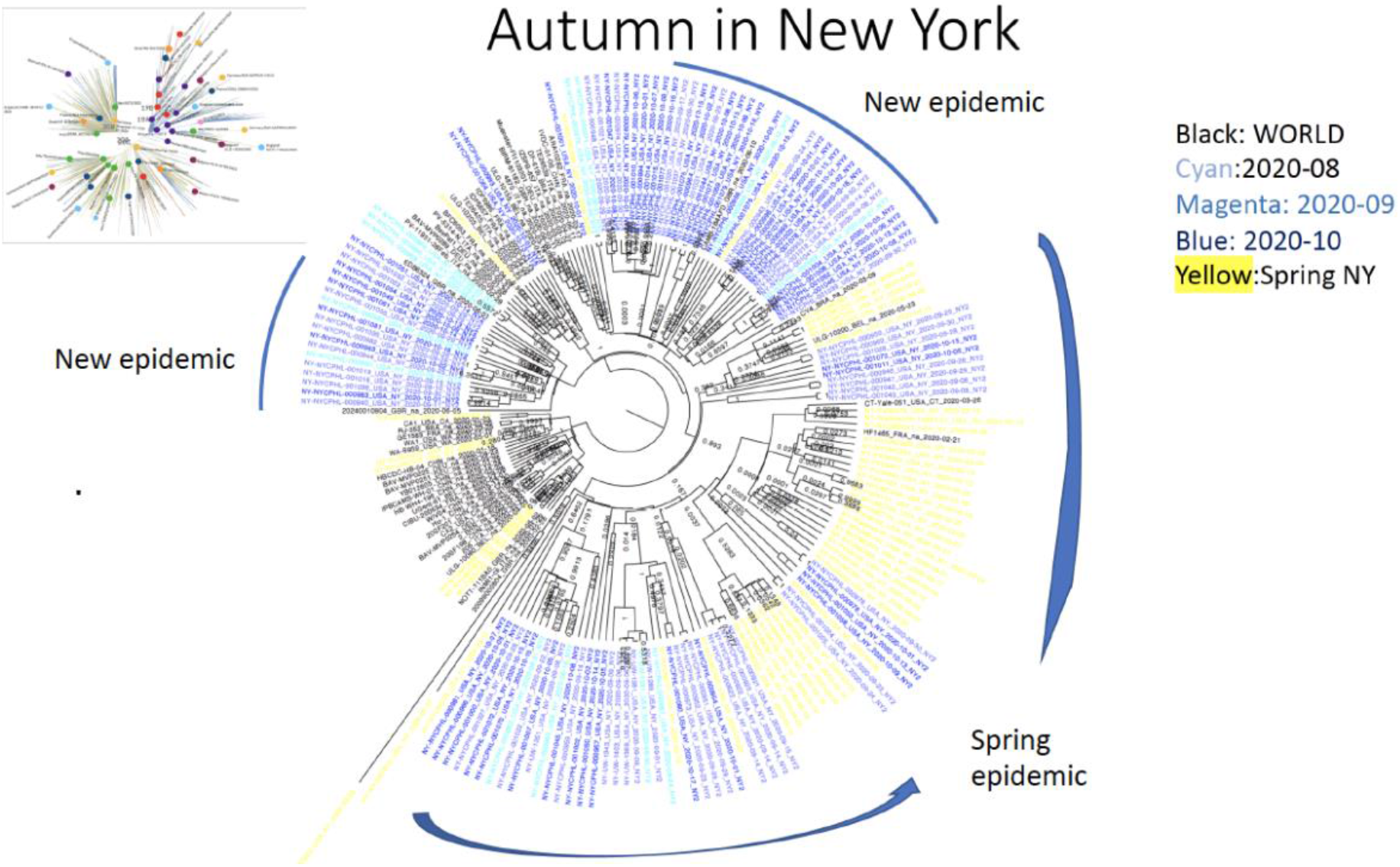
By inferring a phylogenetic tree using sequences from New York that were collected after the 16^th^ of August, together with previous world reference sequences and reference sequences from New York during the first wave, we show that about half of the 2^nd^ wave’s outbreak in New York constitutes a previously unseen outbreak, clustering with reference sequences from Great Britain.

## METHODS

### Data Availability

All data are available in public databases. SARS-CoV2 genomes were retrieved from the GISAID database ^2^. Epidemiological data concerning the daily and total deaths per million individuals have been retrieved from Worldometer ‘worldometers.info/coronavirus/’. Maximum reproduction rates have been retrieved from The Covid Tracking Project “https://covidtracking.com/” and ‘https://rt.live/us/’ ^29^. For the first wave of Covid-19 outbreak in United States we collected a total of 3,133 sequences for the states of New York (NY), Connecticut (CT), Massachusetts (MA), New Jersey (NJ), Pennsylvania (PA), Maryland (MD) and Virginia (VA) that were sampled between 01/29/2020 and 07/05/2020. More specifically, we collected 1505, 353, 418, 45, 112, 178, 522 sequences from each state, respectively. For the second wave of Covid-19 in New York, we collected a total of 112 sequences sampled between 08/01/2020 to 10/18/2020.

#### World reference sequences

For the use of reference sequences representing the global pandemic, we randomly selected 50 sequences spanning Nextstrain lineages 19A, 19B, 20A, 20B and 20C (see figure 2i).

#### State reference sequences

We randomly selected up to 50 reference sequences from each state, prioritizing selection of one sequence per bipartition with higher than 50% posterior probability. Excluding world reference sequences, 50 sequences were selected from NY, CT, MA and VA while 43, 37, 22 were selected from NJ, PA and MD, respectively.

### Phylogenetic Analysis

By retrieving the genomic sequences from GISAID (Supplementary table), we used MAFFT (45) to build multiple sequence alignments for every state based on nucleotide sequence data. Then, using BEAST (31, 32), we performed Bayesian phylogenetic analysis with time constraints based on sampling dates, under a GTR evolutionary model. To determine the appropriate growth models and population size, we tested various growth models including a i) Yule process, ii) exponential growth, iii) logistic growth iv) Bayesian Skyline v) Birth–Death skyline. The BEAST suite also includes multiple software tools that aid in selecting appropriate models and parameters (BEAUti) to infer a phylogenetic tree using Bayesian inference, coalescent theory and speciation with respect to the time of sequence collection. We evaluated the efficacy of these models using Tracer v1.7.1(46). The best model (Yule process) for this data was selected based on the estimated sample size, posterior probabilities, and reports on algorithm convergence.

### Estimating Transitional Connectivity

Using custom scripts, we were able to parse the inferred phylogenetic trees into groups of sequences based on the tree bipartitions. Then, by further parsing the groups in ascending order based on group size (from groups of 2 to X=10), we determined all possible pairs and state connectivity based on dates. For example, pair of sequences {*NY-PV09151_USA_NY_2020-03- 22 and CT-UW-6574_USA_CT_2020-04-03*} would depict an outgoing connectivity between New York (NY) and Connecticut (CT) denoted as NY>CT +1 (see figure 3i). In the manuscript we show results for a strict/conservative approach where pair inconsistencies are dropped, and sequences cannot be considered as incoming twice.

### Maximum Reproduction Rate R_t_

To calculate the maximum reproduction rate Rt, we used the maximum Rt value for each state from ‘https://rt.live/us/’ during the first wave of the pandemic (until August 2020). Rt represents the effective reproduction rate of the virus calculated for each locale. It allows to estimate how many secondary infections are likely to occur from a single infection in a specific area.

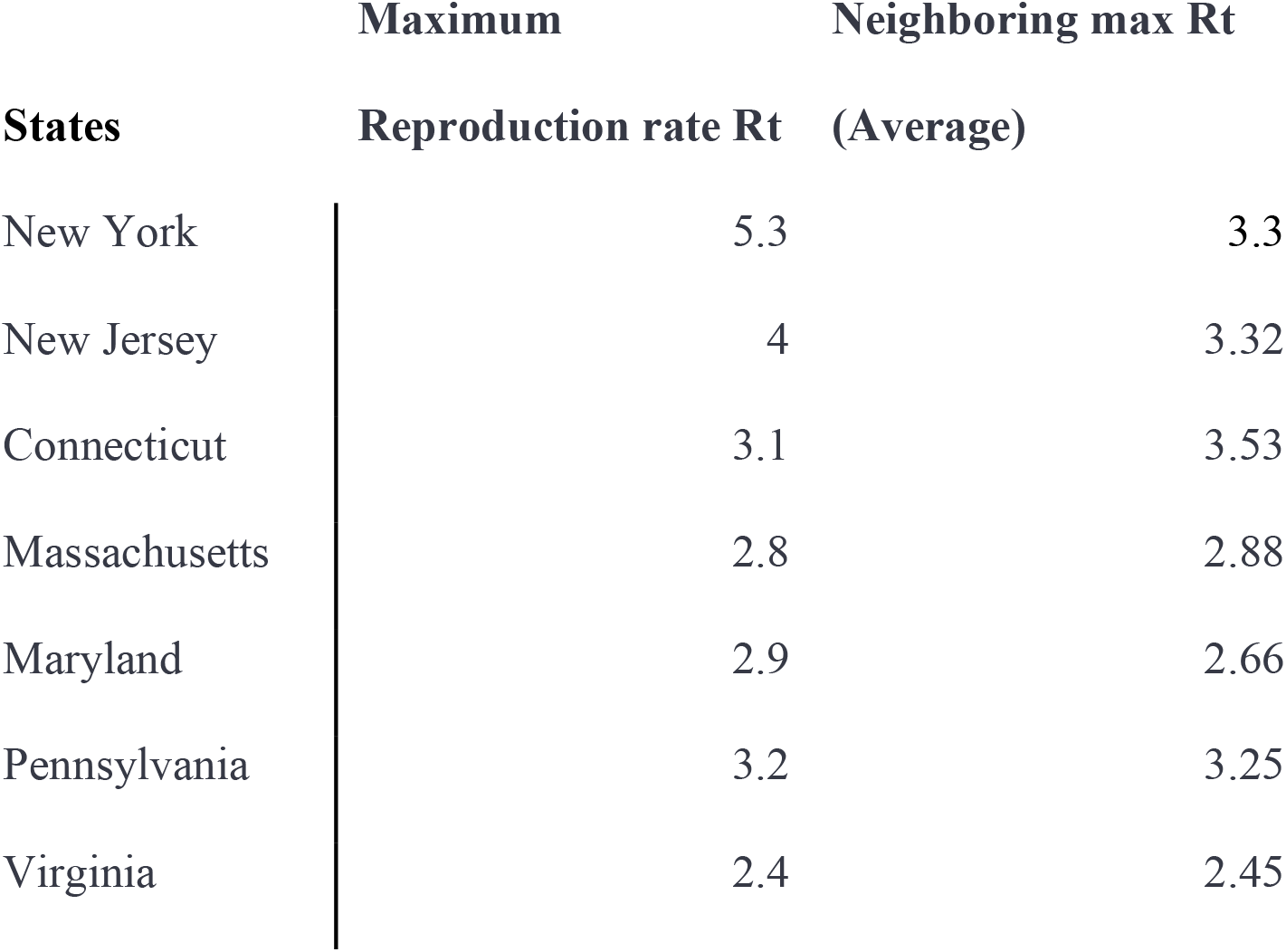
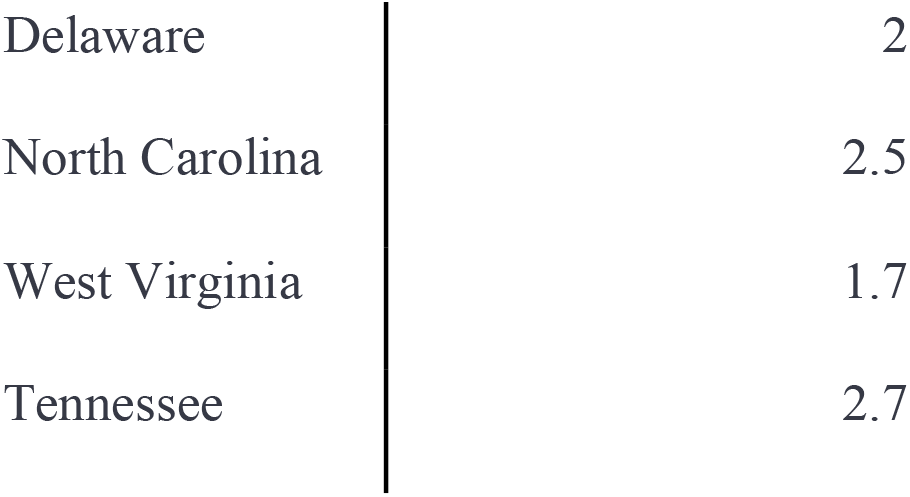

| States | Maximum<br>Reproduction rate $R_t$ | Neighboring max $R_t$<br>(Average) |
| --- | --- | --- |
| New York | 5.3 | 3.3 |
| New Jersey | 4 | 3.32 |
| Connecticut | 3.1 | 3.53 |
| Massachusetts | 2.8 | 2.88 |
| Maryland | 2.9 | 2.66 |
| Pennsylvania | 3.2 | 3.25 |
| Virginia | 2.4 | 2.45 |

|  |  |
| --- | --- |
| Delaware | 2 |
| North Carolina | 2.5 |
| West Virginia | 1.7 |
| Tennessee | 2.7 |

### Urbanization Index

For the Urbanization Index, as an indication of how “urban” a state is, we used the definition and data from 538 (‘https://fivethirtyeight.com/’). FiveThirtyEight’s urbanization index is calculated as the natural logarithm of the average number of people living within a five-mile radius of a given resident.

| State | Urbanism |
| --- | --- |
| New York | 12.56 |
| New Jersey | 12.24 |
| Connecticut | 11.41 |
| Massachusetts | 11.84 |
| Maryland | 11.71 |
| Pennsylvania | 11.15 |
| Virginia | 10.91 |

### Regression Analysis Models

We perform multiple linear regression analyses in order to assess the importance of each factor on the prediction of the per-state death rate. We use data from 7 states (NY, CT, MA, PA, NJ, VA, MD), over a series of 10 timepoints from April 29 to July 23. We regress the per-state death rate (the cumulative ratio of deaths to cases from the earliest date) on either three variables (Transmission rate (R0), Urbanism, Distance from NYC) or six variables (Transmission rate (R0), Urbanism, Distance from NYC, ingoing/outgoing/ingrowing rates per-state). Prior to the analysis, we Z-score all variables (enforcing zero mean and unit covariance). For distance from NYC, we use either the geographic distance between the state’s capital and NYC, or the transition distance as defined below. For each model, we calculate the log-likelihood by fitting a variance parameter to the predicted outputs and using a Gaussian noise model. Hence, we set *σ*_*t*_^2^ = (1/*N*)*Σ*_*i*=1:*N*_(*y*_*it*_ − *β*_***t***_***x***_***it***_)^2^, where *N*is the number of states, *β*_*t*_and *x*_*it*_ are the vectors of coefficients and features associated with state *i* at time *t* respectively, and *y*_*it*_ is the associated death rate. We calculate the log-likelihood at time *t* as *L*_*t*_ = *Σ*_*i*_*log*(*Gauss*(*y*_*it*_ − *β*_***t***_***x***_***it***_; **0**, ***σ***_***t***_)), where *Gauss* is the probability density function of a normal distribution. We then compare the log-likelihood differences of pairs of models over time using the Pearson Correlation Coefficient (differences versus temporal ordering).

### Random Walk Model

We define the transmission distance of a state from NYC as the expected first arrival time at that state of a Markov random walk starting at NYC, using the transition probabilities between states inferred from the phylogenetic analysis. Hence, we set *d*_*ij*_ = *E*(*min*({*t*|*s*_*t*_ = *j*})|*s*_0_ = *i*) for the directed transmission-distance between states i and j (which is not a metric), where *s*_*t*_ = *i* indicates that the state at time *t* in a sampled random walk is *i*, and *E*(.) denotes expectation. To estimate these distances, we run 1000 such random walks for 1000 time-steps and use the empirical mean time of first arrival at each state across samples. As above, we Z-score the resulting distances for each state.

### Mitigation Analysis

In order to break the link between geographically adjacent states *s*_1_ and *s*_2_, we set a reduction factor *r* = 0.1, and update the transmission probabilities as: *P*′(*s*_*b*_|*s*_*a*_) = *r* · *P*(*s*_*b*_|*s*_*a*_), and *P*′(*s*_*a*_|*s*_*a*_) = *P*(*s*_*a*_|*s*_*a*_) + (1 − *r*) · *P*(*s*_*b*_|*s*_*a*_), where *P*′(*s*_*a*_|*s*_*b*_) is the updated transition probability between states *s*_*a*_ and *s*_*b*_. We make such updates for *a* = 1, *b* = 2 and *a* = 2, *b* = 1 simultaneously, hence breaking the link in both directions. We then recalculate the distances 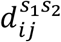, i.e. the distance between states i and j, given the link between *s*_1_ and *s*_2_ has been broken. We then use these to estimate the overall predicted reduction in the death-rate given the break as: 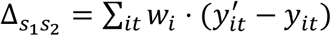, where w_i_ is a weighting factor proportional to the population of state *i* (and ∑_*i*_ *w*_*i*_ = 1), and 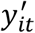 is the predicted death-rate for state i at time t when 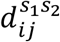 is substituted for *d*_*ij*_ in the predictive model from the Regression analysis.

## DISCUSSION

Previous studies have provided an important historic view of travel history ^8,9,11–14,30^ and viral spread of Covid-19^6,11,12,15^ using genetic variability. Others, data driven, have modeled the spread of the virus and effectiveness of government interventions ^20,27,28^. So far, the only acclaimed and efficient non-pharmaceutical interventions in our arsenal are forms of regional lockdowns ^19,22,26,31,32^, while other strategies relying on ‘herd immunity’ have also been suggested and disputed ^23–25^.

Here, we used SARS-CoV2 genomes to determine regional connectivity in a case study of 7 states, where New York acted as an initial hotspot. By combining epidemiological demographic and genetic information, we used four regression models to evaluate the importance of different factors that contribute to outbreak severity throughout the first viral wave.

Our results can explain the discordance between regions and strategies, especially between the first and second pandemic waves. For example, states within distance from hotspots are able to deal with a milder initial outbreak, before the virus establishes at a later timepoint, depending on transitional distance (i.e., the speed of the wave) and regional connectivity. Similarly, states with lower connectivity (e.g., naturally or physically isolated regions) can be more efficient in battling the viral spread, as they deal with reduced viral wave and incoming infections. This also suggests that reducing incoming transmission routes (through pharmaceutical or non-pharmaceutical interventions) can have a significant effect in addition to local mitigation strategies such as lockdowns. This does not necessarily mean complete isolation, but rather a blockade on transmission routes with high connectivity. However, our results also suggest that states deciding to follow less stringent mitigation strategies are also largely responsible for their outgoing viral connectivity, affecting neighboring regions, while often taking advantage of the low incoming connectivity resulting from neighboring lockdowns in return.

By deriving genetic connectivity between regions using genomic information, we combined genetic information with demographic and epidemiological data to create a model and a proxy for the flow of the viral wave in order to study factors that temporally contribute to the severity of local outbreaks throughout the pandemic. Then we used this model to consider the outcome of selective intervention strategies using geographic blockades. Overall, our results suggest that unified mitigation strategies are more efficient in tackling a pandemic, while also providing a framework within which to pursue these strategies. Our framework can be implemented for both pharmaceutical (e.g vaccination) or non-pharmaceutical interventions (e.g., lockdowns, blockades).

## Data Availability

All data are available in public databases.

## Author Contributions

L.S. conceived of the project, designed, developed, performed, and analyzed experiments. J.W. developed and performed the regression and random walk models. H.C. performed the phylogenetic analysis. A.C. performed the regression and random walk models. L.S. drafted the paper. L.S., J.W., and M.G. wrote the paper. All authors read and approved the final paper.

## BIBLIOGRAPHY

1. Sayers, E. W. et al. GenBank. Nucleic Acids Res. (2020). doi:10.1093/nar/gkz956

2. GISAID. GISAID Initiative. Adv. Virus Res. (2020).

3. Hadfield, J. et al. NextStrain: Real-time tracking of pathogen evolution. Bioinformatics (2018). doi:10.1093/bioinformatics/bty407

4. Leitner, T. et al. HIV Sequence Compendium 2008 Los Alamos HIV Sequence Database. HIV Seq. Compend. (2008).

5. Kuiken, C., Hraber, P., Thurmond, J. & Yusim, K. The hepatitis C sequence database in Los Alamos. Nucleic Acids Res. (2008). doi:10.1093/nar/gkm962

6. Candido, D. S. et al. Evolution and epidemic spread of SARS-CoV-2 in Brazil. Science (80-.). (2020). doi:10.1126/SCIENCE.ABD2161

7. Isabel, S. et al. Evolutionary and structural analyses of SARS-CoV-2 D614G spike protein mutation now documented worldwide. Sci. Rep. (2020). doi:10.1038/s41598-020-70827-z

8. Lemey, P. et al. Accommodating individual travel history and unsampled diversity in Bayesian phylogeographic inference of SARS-CoV-2. Nat. Commun. (2020). doi:10.1038/s41467-020-18877-9

9. Seemann, T. et al. Tracking the COVID-19 pandemic in Australia using genomics. Nat. Commun. (2020). doi:10.1038/s41467-020-18314-x

10. Deng, X. et al. Genomic surveillance reveals multiple introductions of SARS-CoV-2 into Northern California. Science (80-.). (2020). doi:10.1126/science.abb9263

11. Jorden, M. A. et al. Evidence for Limited Early Spread of COVID-19 Within the United States, January–February 2020. MMWR. Morb. Mortal. Wkly. Rep. (2020). doi:10.15585/mmwr.mm6922e1

12. Fauver, J. R. et al. Coast-to-Coast Spread of SARS-CoV-2 during the Early Epidemic in the United States. Cell (2020). doi:10.1016/j.cell.2020.04.021

13. Bedford, T. et al. Cryptic transmission of SARS-CoV-2 in Washington State. Science (80-.). (2020). doi:10.1101/2020.04.02.20051417

14. Worobey, M. et al. The emergence of SARS-CoV-2 in Europe and North America. Science (80-.). (2020). doi:10.1126/SCIENCE.ABC8169

15. Xu, B. et al. Epidemiological data from the COVID-19 outbreak, real-time case information. Sci. Data (2020). doi:10.1038/s41597-020-0448-0

16. Wynants, L. et al. Prediction models for diagnosis and prognosis of covid-19: Systematic review and critical appraisal. BMJ (2020). doi:10.1136/bmj.m1328

17. Weinberger, D. M. et al. Estimation of Excess Deaths Associated with the COVID-19 Pandemic in the United States, March to May 2020. JAMA Intern. Med. (2020). doi:10.1001/jamainternmed.2020.3391

18. Ioannidis, J. P. A., Axfors, C. & Contopoulos-Ioannidis, D. G. Population-level COVID- 19 mortality risk for non-elderly individuals overall and for non-elderly individuals without underlying diseases in pandemic epicenters. Environ. Res. (2020). doi:10.1016/j.envres.2020.109890

19. Eubank, S. et al. Commentary on Ferguson, et al., “Impact of Non-pharmaceutical Interventions (NPIs) to Reduce COVID-19 Mortality and Healthcare Demand”. Bull. Math. Biol. (2020). doi:10.1007/s11538-020-00726-x

20. Cacciapaglia, G., Cot, C. & Sannino, F. Second wave COVID-19 pandemics in Europe: a temporal playbook. Sci. Rep. (2020). doi:10.1038/s41598-020-72611-5

21. Reiner, R. C. et al. Modeling COVID-19 scenarios for the United States. Nat. Med. (2020). doi:10.1038/s41591-020-1132-9

22. Ferguson, N. et al. Report 9 - Impact of non-pharmaceutical interventions (NPIs) to reduce COVID-19 mortality and healthcare demand Faculty of Medicine Imperial College London. Imp. Coll. COVID Response Team (2020).

23. Jung, F., Krieger, V., Hufert, F. T. & Küpper, J. H. Herd immunity or suppression strategy to combat COVID-19. Clin. Hemorheol. Microcirc. (2020). doi:10.3233/CH-209006

24. Orlowski, E. J. W. & Goldsmith, D. J. A. Four months into the COVID-19 pandemic, Sweden’s prized herd immunity is nowhere in sight. J. R. Soc. Med. (2020). doi:10.1177/0141076820945282

25. Aschwanden, C. The false promise of herd immunity for COVID-19. Nature (2020). doi:10.1038/d41586-020-02948-4

26. Farsalinos, K. et al. Improved strategies to counter the COVID-19 pandemic: Lockdowns vs. primary and community healthcare. Toxicol. Reports (2021). doi:10.1016/j.toxrep.2020.12.001

27. Fang, Y., Nie, Y. & Penny, M. Transmission dynamics of the COVID-19 outbreak and effectiveness of government interventions: A data-driven analysis. J. Med. Virol. (2020). doi:10.1002/jmv.25750

28. Brauner, J. M. et al. Inferring the effectiveness of government interventions against COVID-19. Science (80-.). (2020). doi:10.1126/science.abd9338

29. Kevin Systrom, T. V. and M. K. Rt.live. (2020).

30. Mbuvha, R. & Marwala, T. Bayesian inference of COVID-19 spreading rates in South Africa. PLoS One (2020). doi:10.1371/journal.pone.0237126

31. Panovska-Griffiths, J. et al. Determining the optimal strategy for reopening schools, the impact of test and trace interventions, and the risk of occurrence of a second COVID-19 epidemic wave in the UK: a modelling study. Lancet Child Adolesc. Heal. (2020). doi:10.1016/S2352-4642(20)30250-9

32. Stefana, A., Youngstrom, E. A., Hopwood, C. J. & Dakanalis, A. The COVID- 19 pandemic brings a second wave of social isolation and disrupted services. European Archives of Psychiatry and Clinical Neuroscience (2020). doi:10.1007/s00406-020-01137-8

