## Supplementary figures for "Genetic determination of regional connectivity in modelling the spread of COVID-19 outbreak for improved mitigation strategies"

**Figure S1-7.** Individual state trees. In Yellow, we show the respective state sequences, while in black the world reference sequences. Branch labels represent posterior probability.

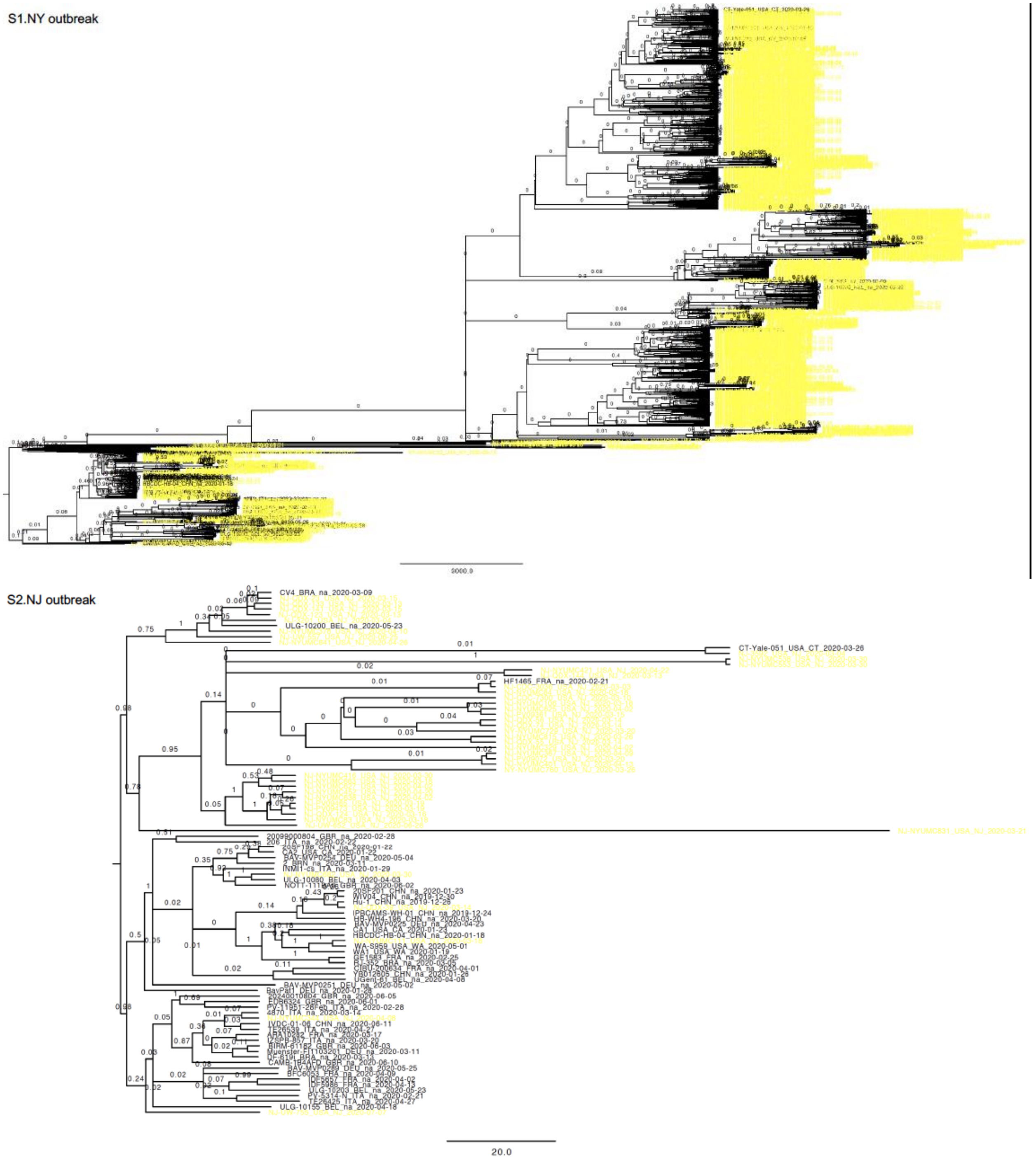

S3. CT outbreak

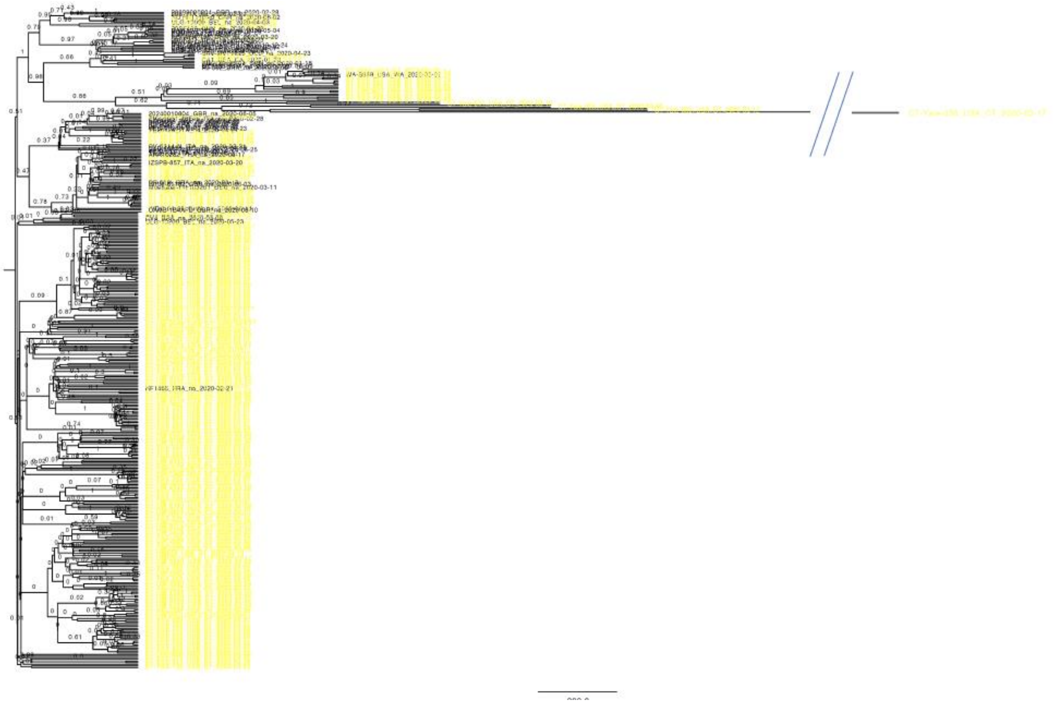

S4. MA outbreak

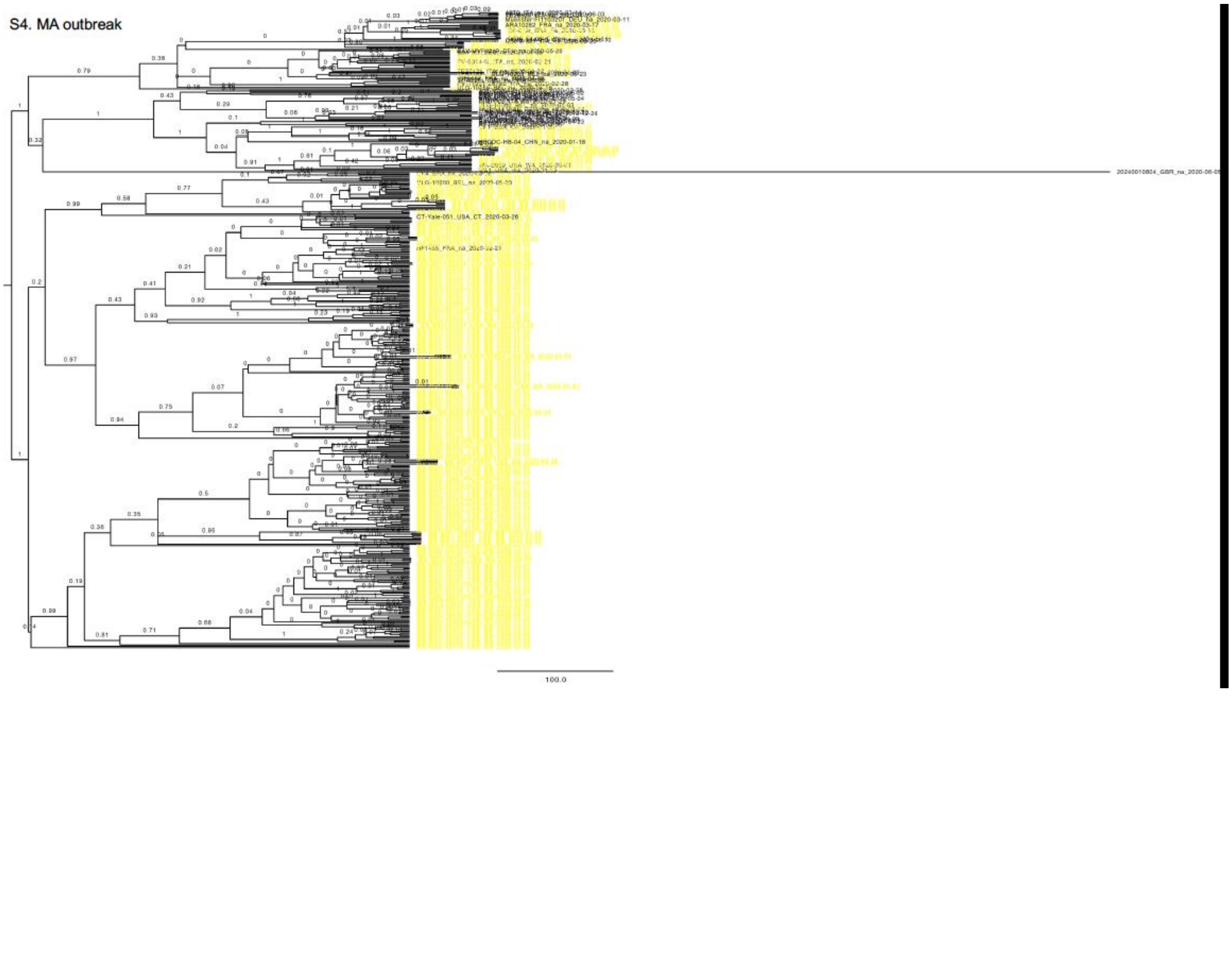

S5. PA outbreak

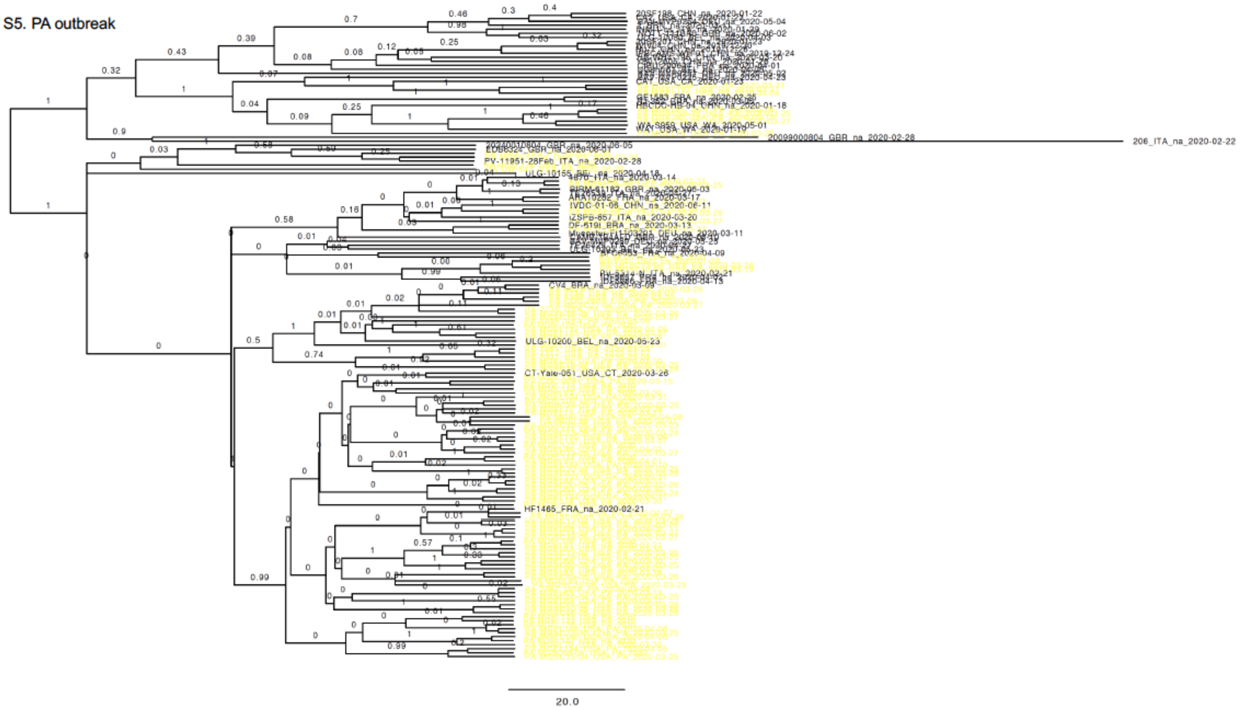


S6. MD outbreak

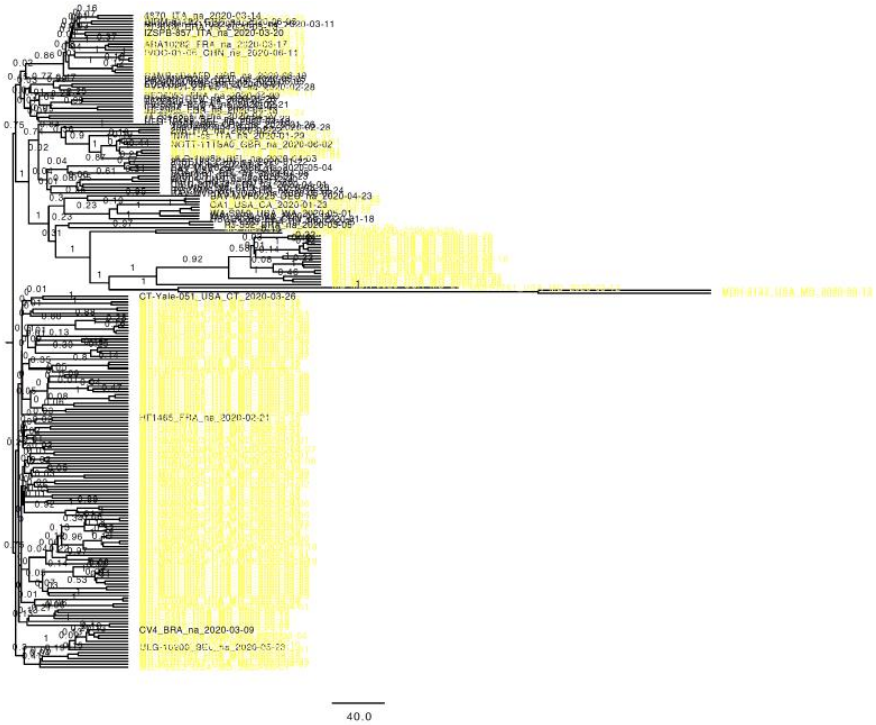

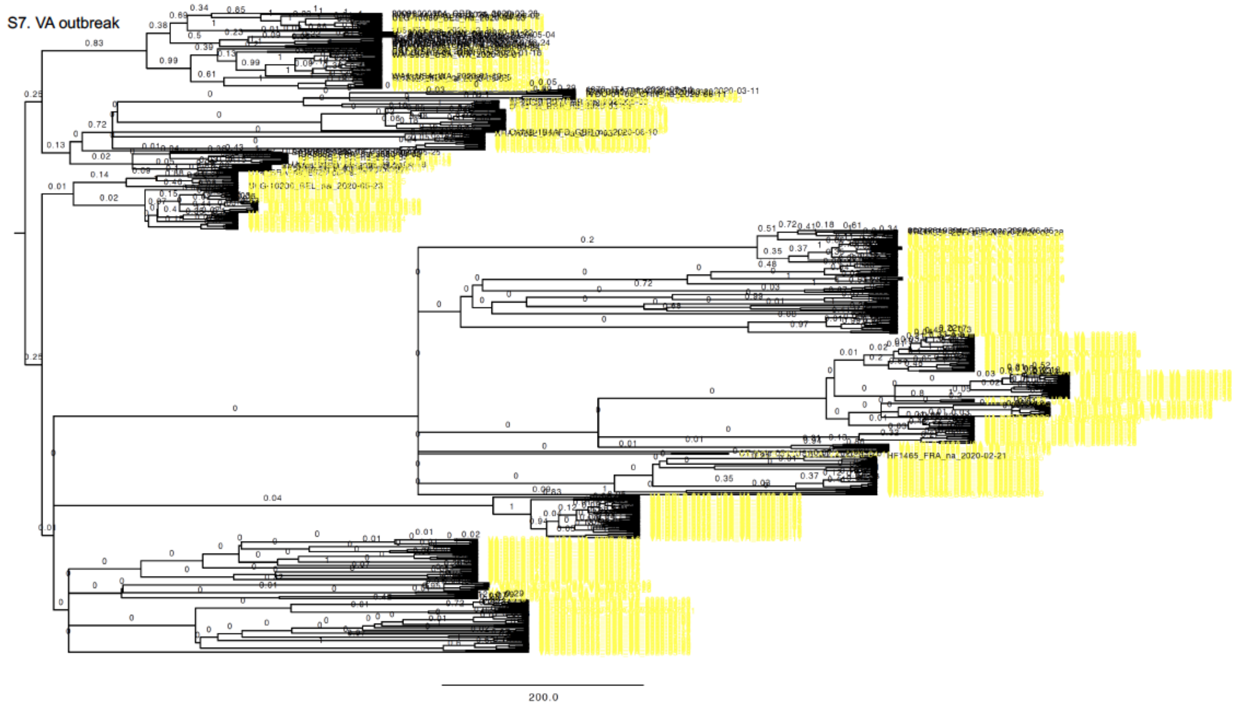


**Figure S8.** Temporal Likelihood difference for nested models [4ii vs 4i, solid line] and [4iv vs 4iii, dotted line]; (p=0.0003, 0.0273 respectively on 2-sided t-test)

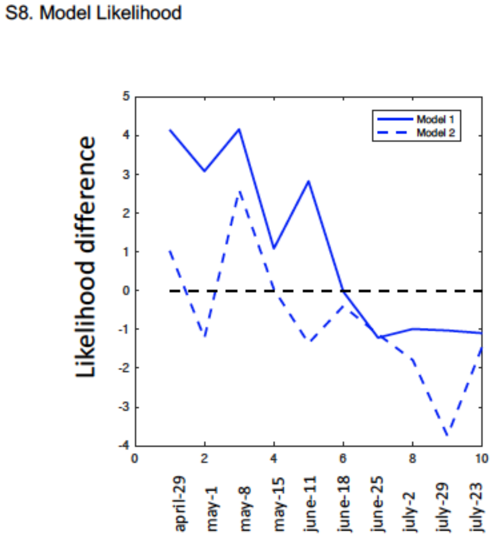


**Figure S9.** Daily deaths for 7 states as adapted from ‘https://www.worldometers.info/coronavirus/’. In states distant from NYC, we observe a lag-time between $T_0$ and the peak of daily deaths. In the extreme case of VA, there is not a clear peak during the first wave.

### S9. Daily deaths

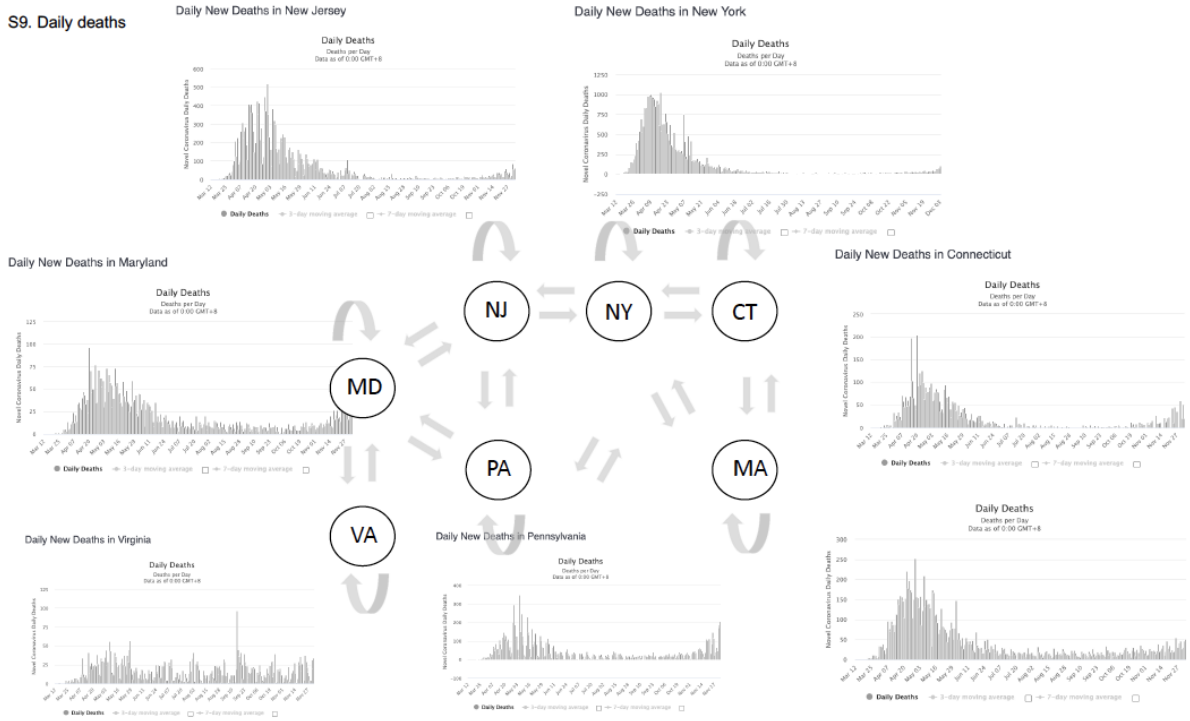
